# Movement history influences pendulum test kinematics in children with spastic cerebral palsy

**DOI:** 10.1101/2020.04.07.20048314

**Authors:** Willaert Jente, Kaat Desloovere, Anja Van Campenhout, Lena H. Ting, Friedl De Groote

## Abstract

The pendulum test assesses quadriceps spasticity by dropping the lower leg of a relaxed patient from the horizontal position and observing limb movement. The first swing excursion decreases with increasing spasticity severity. Our recent simulation study suggests that the reduced initial swing results from muscle short-range stiffness and its interaction with reflex hyper-excitability. Short-range stiffness emerges from the thixotropic behavior of muscles where fiber stiffness upon stretch increases when the muscle is held isometric. Fiber stiffness might thus be higher during the first swing of the pendulum test than during consecutive swings. In addition, it has recently been suggested that muscle spindle firing reflects fiber force rather than velocity and therefore, reflex activity might depend on fiber stiffness. If this hypothesized mechanism is true, we expect to observe larger first swing excursions and reduced reflex muscle activity when the leg is moved rather than kept isometric before release, especially in patients with increased reflex activity. We performed the pendulum test in 15 children with cerebral palsy (CP) and 15 age-matched typically developing (TD) children in two conditions. In the hold condition, the leg was kept isometric in the extended position before release. In the movement condition, the leg was moved up and down before release to reduce the contribution of short-range stiffness. Knee kinematics and muscle activity were recorded. Moving the leg before release increased first swing excursion (p < 0.001) and this increase was larger in children with CP (21°) than in TD children (8°) (p < 0.005). In addition, pre-movement delayed reflex onset by 87 ms (p < 0.05) and reduced reflex activity as assessed through the area under the curve of rectus femoris electromyography (p < 0.05) in children with CP. The movement history dependence of pendulum kinematics and reflex activity supports our hypothesis that muscle short-range stiffness and its interaction with reflex hyper-excitability contribute to joint hyper-resistance in spastic CP. Our results have implications for standardizing movement history in clinical tests of spasticity and for understanding the role of spasticity in functional movements, where movement history differs from movement history in clinical tests.

## Introduction

Spasticity is a common symptom in patients with cerebral palsy (CP) or stroke, but the underlying mechanisms are poorly understood. Spasticity is traditionally defined as ‘a velocity-dependent increase in tonic stretch reflex resulting from hyper-excitability of the stretch reflex’ (Lance JW 1980). Clinical tests of spasticity, such as the Modified Ashworth Scale (MAS) or Tardieu Scale, assess the resistance of the joint to imposed movement at different speeds. In this paper, we will use the terms spasticity and joint hyper-resistance against movement interchangeable. These clinical tests give little insight in the underlying mechanisms of increased resistance to movement, which might explain why the contribution of spasticity to walking impairments is only poorly understood (Papageorgiou et al. 2019). There is growing consensus that it is important to distinguish different contributions to joint hyper-resistance, i.e. non-neural originating from passive tissue properties, and neural originating from background muscle activity and stretch hyperreflexia (van den Noort et al. 2017). Our recent simulation study of the pendulum test of spasticity suggests that background muscle activity interacts with stretch hyperreflexia through muscle short-range stiffness that is dependent on movement history (De Groote et al. 2018). Hence, interactions between neural contributions and history-dependent muscle mechanics might be a crucial determinant of joint hyper-resistance to stretch in movement impairments. Yet, we currently lack experimental evidence for the movement history dependence of joint hyper-resistance.

During the pendulum test, an examiner drops the leg of a seated and relaxed patient from the horizontal position and the lower leg then swings freely under the influence of gravity (Fowler et al. 2000), (figure 1a). Upon release of the leg, the quadriceps muscles are stretched. In healthy individuals, the lower leg behaves like a damped pendulum. With increasing levels of spasticity, (1) the amplitude of the first swing excursion decreases, (2) the number of oscillations decreases, and (3) the position of the lower leg when it comes to rest, described by the resting angle, is less vertical (figure 1b). The first swing excursion has been found to be the strongest predictor of spasticity severity (Fowler et al. 2000; Szopa et al. 2014).

**Figure 1:**
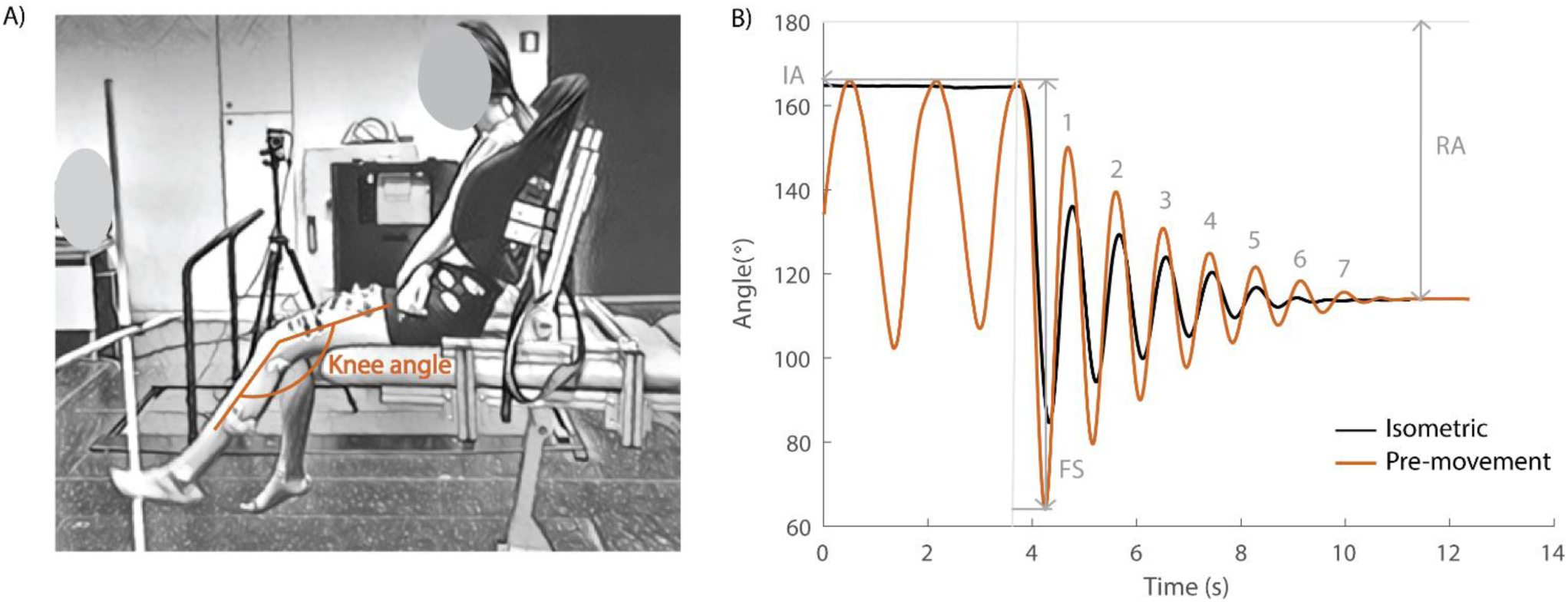
Pendulum test set-up and definition of kinematic outcome parameters. A) Subject leaned against a custom-made backrest in the seated position. The knee angle of the swinging leg is indicated in orange. B) Knee angle trajectory during an isometric (HR) trial in black and a pre-movement (MR) trial in orange in the sitting position with indication of kinematic outcome parameters. IA = initial angle; FS = first swing excursion; RA = resting angle; numbers indicate the number of oscillations for the orange trajectory. The vertical grey line indicates release of the leg.

We performed a simulation study to test the neuro-mechanical mechanisms underlying altered pendulum test kinematics in children with spastic CP and found that modeling muscle tone, i.e. background muscle activity, and transient short-range stiffness were crucial to simulate decreased first swing excursion and non-vertical resting angle (De Groote et al. 2018). Short-range stiffness emerges from the thixotropic behavior of muscles where fiber stiffness upon stretch depends on movement history with stiffness increasing when the muscle is held at a constant length (Campbell KS and Lakie M 1998). When a muscle fiber is consecutively stretched and released multiple times, the increase in force is significantly higher during the first stretch than during the later stretches. Short-range stiffness leads to a sharp and rapid increase in force upon stretch of an isometric muscle until a force plateau is reached at the critical stretch of the muscle (Campbell and Moss 2002). Both the initial rate of force development and force plateau depend on the level of isometric force before the stretch (Campbell and Moss 2002), and therefore on the level of background muscle activity and through the muscle’s force-length relationship on isometric fiber length (Gordon and Ridgway 1987; Herzog and ter Keurs 1988). The isometric period before releasing the leg in the pendulum test and the high prevalence of increased background muscle activity in CP (Freeman Miller 2005) might therefore lead to large contributions from short-range stiffness during the first swing limiting leg excursion. In addition, background muscle activity of the quadriceps might also contribute to the non-vertical resting angle.

Furthermore, our simulation study suggested an interaction between muscle short-range stiffness and reflex activity. The reduction in the number of oscillations was best reproduced by simulating stretch reflex activity in terms of force, and not velocity, feedback (De Groote et al. 2018). Our model of reflex activity was based on prior experimental work in cats in which sensory signals from muscle spindles are transiently increased in response to muscle stretch after being held at a constant length. Spindle firing was strongly correlated with muscle short-range stiffness force and force rate, i.e. yank, (Blum et al. 2017; Lin et al. 2019). Similarly, short-range stiffness might interact with hyperactive stretch reflexes during the pendulum test in individuals with spastic CP to reduce first swing excursion and thereby the number of subsequent oscillations.

Here, we tested the hypothesis that short-range stiffness due to increased muscle tone and its interaction with reflexes contribute to abnormal pendulum test kinematics in spastic CP. To this aim, we performed the pendulum test in different conditions that were designed to alter the contribution of short-range stiffness in children with spastic CP and age-matched typically developing (TD) children. In the isometric condition, the leg was held still for at least five seconds prior to release whereas in the pre-movement condition, the leg was moved up and down prior to release to decrease the contribution of short-range stiffness. If short-range stiffness indeed contributes to abnormal pendulum test kinematics in spastic CP, pre-movement should normalize pendulum test kinematics by increasing the first swing excursion and number of oscillations. However, pre-movement is not expected to alter the resting angle, which is assumed to be determined by muscle tone and passive muscle properties, and thus not altered by movement history. In addition, if short-range stiffness indeed interacts with reflex activity, pre-movement of the leg should reduce reflex activity (Blum et al. 2017). More precisely, we expect the slower build-up of force, due to decreased short-range stiffness in the pre-movement condition, to reduce and delay the peak in reflex activity. Finally, we tested the dependency of joint hyper-resistance on active muscle force by altering the pose of the subjects from sitting to supine, which increases rectus femoris length. The muscle’s force-length relationship predicts a decrease in active force due to muscle tone and an increase in passive force when a muscle is stretched beyond its optimal length (Gordon and Ridgway 1987; Herzog and ter Keurs 1988). As short-range stiffness is proportional to active force, we expect the effect of pre-movement to be smaller in the supine than in the sitting position. Since TD children have lower muscle tone and reflexes than children with CP when relaxed (Dietz and Sinkjaer 2007; Freeman Miller 2005), we expect a smaller effect of pre-movement in TD children.

## Materials and Methods

### Subjects

Thirty children (20 boys, 10 girls) participated in this study. Fifteen children with spastic CP were recruited through the clinical motion analysis laboratory at the University Hospital of Leuven (Belgium). Measurements were planned following the clinical gait analysis that was part of the routine clinical follow-up for children with CP. All patients were diagnosed as having spastic CP confirmed by a neuro-paediatrician. Following inclusion criteria were used: (1) age between 5 years and 17 years; (2) Gross Motor Function Classification Scale (GMFCS) level I - III; (3) no orthopedic or neurological surgery in the previous year; and (4) no Botulinum toxin injections at least 6 months before the measurements of this study. The CP group consisted of children with unilateral (N = 3) and bilateral (N = 12) spasticity. Eight of them had GMFCS level I, five GMFCS level II and two GMFCS level III. Four children had spasticity in both the quadriceps and hamstring muscles, nine children had spasticity only in the hamstring muscles and two children had no hamstrings or quadriceps spasticity as measured by the Modified Ashworth Scale (table S1). Fifteen TD children, which were recruited through colleagues and friends, volunteered as controls. All children (CP and TD) were aged between 5 and 14 years old and groups were age matched (table 1). For children with CP, the most affected leg was tested, while for TD children one leg was randomly selected.

**Table 1:** Demographic data of the participants

|  | CP |  | TD |  |
| --- | --- | --- | --- | --- |
|  | Mean | SD | Mean | SD |
| **Female/male** | 3/12 |  | 7/8 |  |
| **Age (years)** | 9.7 | 2.8 | 10.0 | 2.5 |
| **Length (cm)** | 138 | 18 | 142 | 15 |
| **Weight (kg)** | 33.4 | 12.1 | 33.6 | 10.0 |

### Materials

Children with spastic CP were tested in the clinical motion analysis laboratory at the University hospital of Leuven (Belgium). TD children were tested in the Movement and Posture analysis Laboratory Leuven (MALL, Belgium). Both labs are equipped with a Vicon camera system (Vicon, Oxford Metrics, UK, 100 Hz). Kinematics were measured by placing reflective markers that could be detected by infrared cameras on specific anatomic landmarks on the participant (for details on marker placement, see figure S1). Muscle activity was measured simultaneously by a wireless surface electromyography system (sEMG) (CP: Wave Wireless EMG, Biometrics, United Kingdom, 1000 Hz; TD: ZeroWire EMG Aurion, Cometa, Italy, 1000 Hz) with silver-chloride, pre-gelled bipolar electrodes (CP: Nutrode, Xsanatec, Belgium; TD: Ambu Blue Sensor, Ballerup, Denmark). Muscle activity was measured for four lower limb muscles (m. rectus femoris, m. vastus lateralis, m. semitendinosus and m. biceps femoris). All electrodes were placed according to SENIAM guidelines (Hermens et al. 2000). A custom-made backrest was built to provide back support during the pendulum test. This chair allowed us to control and standardize the hip angle between subjects and trials. The pendulum test was performed in two positions: a sitting and supine position. On average, the hip was 28° (±11°) into flexion for the sitting position and 8° (± 8°) into extension for the supine position.

### Protocol

The protocol was approved by the Ethical Committee of UZ Leuven/KU Leuven (s61641). A legal representative of the participant signed the informed consent and participants older than 12 years signed informed assent in accordance with the Declaration of Helsinki before measurements were started. For every subject, age and anthropometrics (length and weight) were collected and sEMG electrodes and markers were placed. Children with CP had a clinical examination and clinical gait analysis before the pendulum test was performed. Rest in between measurements was provided when needed.

During the pendulum test, the child was asked to sit relaxed on the table with back support (figure 1a). The examiner extended the leg to a horizontal position (initial angle) and then dropped the leg. The leg was then allowed to swing freely under the influence of gravity until it came to rest. Kinematic trajectories and sEMG signals were recorded during the test. In each position (sit and supine), the pendulum test was performed in two different conditions; (1) with the leg isometric prior to release (hold and release - HR) and (2) with pre-movement of the leg prior to release (movement and release (MR)). For the sitting position, the back support was used to assure a relaxed position. By flexing the hip, the possible effect of hamstring muscle tightness was minimized. In the supine position, no backrest was used and children were lying in their natural supine position on the table. During the HR condition, the leg was held as close to horizontal as soft tissues allowed for at least five seconds prior to release (presented in all figures in black (sit) or grey (supine)). In the MR condition, small oscillatory movements away from and back to the horizontal position were made with the leg prior to release (presented in all figures in orange (sit) or yellow (supine)). Every condition (HR/MR) was repeated five times or until at least three trials during which the subject was relaxed were obtained with the subject both sitting and supine. Relaxation of the subjects was assessed during the test by real-time visual inspection of the sEMG signals. All trials and subjects were measured by the same examiner.

### Data processing and analysis

Marker trajectories from Nexus (Version 2.8.5, Oxford Metrics, United Kingdom) were processed using OpenSim 3.3 to calculate knee joint angles (Delp et al. 2007; Seth et al. 2018). Key kinematic parameters, first swing excursion (FS), number of oscillations (NO) and resting angle (RA) (figure 1b), were calculated using Matlab (R2018b, Mathworks, United States of America). Raw sEMG data were band-pass filtered using a fourth order Butterworth filter between 10 and 450 Hz followed by signal rectification. Finally, a fourth order Butterworth low-pass filter with 20 Hz cut-off was applied (De Luca et al. 2010).

Key kinematic parameters were defined following Fowler et al. (Fowler et al. 2000) (Figure 1b). First swing excursion was calculated as the difference between the initial knee joint angle (180° corresponds to full extension) and the knee joint angle at the first reversal of the swinging limb. The initial angle was the position at which the examiner released the participant’s heel. We defined release onset as the first frame with angular velocity of the knee < -0.01 rad/s. The number of oscillations was determined by counting the maxima of the sinusoidal waves produced by the swinging limb after the heel was released. The criterion for each oscillation was a displacement of at least 3° towards extension. The resting angle was the lower leg orientation relative to horizontal when the lower leg came to rest after the oscillatory movement. The initial angle was standardized during data collection within each position and subject by placing a visual reference point next to the subject. There was no difference in initial angle observed between the HR and MR condition in the sitting position. However, the leg was on average 1.6° (max: 7.1°) and 2. 1° (max: 6.4°) closer to horizontal in respectively children with CP and TD children in the MR condition with respect to the HR condition in the supine position.

Reflex activity was assessed using three outcome parameters: (1) the occurrence of reflex activity, (2) reflex onset and (3) reflex magnitude. The occurrence of reflex activity was expressed as the percentage of trials that showed reflex activity. The presence of muscle reflex activity was determined by evaluating whether sEMG after release of the leg exceeded average baseline muscle activation in the two seconds prior to release by eight times the standard deviation. We chose a threshold of eight times standard deviation, since lower thresholds resulted in a lot of false positive cases based on visual inspection of the sEMG signal. When reflex activity was observed, the onset was defined as the first frame where muscle activity exceeded baseline muscle activation by four times the standard deviation. We could use a lower threshold here as the higher threshold used for detection prevented false detection of reflex activity. The change in reflex magnitude between conditions was assessed based on the area under the curve (AUC) of the baseline subtracted sEMG signal from release onset until 500 ms after release onset. Since the magnitude of sEMG signals cannot be compared between subjects, we used a relative measure of the change in AUC between conditions. The relative difference in AUC between the HR and MR conditions was calculated as the ratio of the difference between the AUC of the HR and MR condition, and the AUC of the HR condition. For subject 4 of the CP group, no sEMG signals were collected during the sitting position for the HR condition due to technical issues and this patient was therefore omitted from sEMG analyses in this condition.

All trials were checked for voluntary activity by visual inspection of the sEMG and kinematic signals. Trials were excluded from further analysis based on sEMG (1) when muscle activity was higher prior to than after release of the leg or (2) when the increase in muscle activity after release of the leg could not be attributed to reflexes based on the criterion that reflexes occurred during muscle stretch and not shortening. Trials were excluded based on kinematic signals (1) when the amplitude of the oscillations increased over time (not taking into account the first swing amplitude) or (2) when the leg came to an abrupt stop. When the child was relaxed during the beginning of the trial but not afterwards according to the above criteria, we still included the trial to analyze outcome parameters that were not affected by voluntary activation in later parts of the trial (e.g. first swing excursion).

### Statistical analysis

Key kinematic parameters (first swing excursion, number of oscillations, and resting angle) and reflex outcomes (occurrence of reflex activity, onset of reflex activity, and reflex magnitude between conditions) were averaged across trials within each condition and subject. Normality was tested using the Shapiro Wilk test. An unpaired t-test was used to compare key kinematic and reflex outcomes between children with CP and TD children. A paired t-test was used to compare outcome parameters between the HR and MR conditions to investigate the influence of movement history. Further, a paired t-test was used to compare outcome parameters between the sitting and supine positions to investigate the influence of position.

We also assessed correlations between outcome parameters that are dependent on the same underlying deficits according to the hypothesized mechanism. First, we assessed the correlation between first swing excursion and resting angle in the isometric condition as both reduced first swing excursion and less vertical resting angle could be explained by increased muscle tone in our model. Second, we calculated the correlation between first swing excursion and reflex activity in the isometric condition as both reduced first swing excursion and increased reflex activity resulted from short-range stiffness in our model. All correlations were calculated using Pearson’s correlations. An alpha level of 0.05 was chosen as cut-off for all statistical comparisons. All statistical analyses were performed using IBM SPSS software version 22 (SPSS Inc., Chicago, IL, United States of America).

## Results

### Information on included trials

All recruited children (15 CP and 15 TD) finished the full test battery. After excluding trials with voluntary muscle activity prior to or during the first swing (see method section), 274 trials (mean: 4.56 trials per condition) were retained for children with CP and 258 (mean: 4.3 trials per condition) trials were retained for TD children to analyze first swing excursion (range 2-8 trials per subject and condition) (table 2). For number of oscillations and resting angle, 75.9% (CP) and 74.8% (TD) of these trials (range 1-7 trials) were used. To analyze reflex activity, 97.8% (CP) and 99.6% (TD) of these trials (range 2-7 trials) were used (table 2).

**Table 2:** Information on the number of included trials

|  |  |  | CP |  |  | TD |  |  |
| --- | --- | --- | --- | --- | --- | --- | --- | --- |
|  |  |  | # | mean | range | # | mean | range |
| IA | Sit | HR | 69 | 4.6 | 3-8 | 64 | 4.3 | 3-5 |
|  |  | MR | 65 | 4.3 | 3-5 | 62 | 4.3 | 2-5 |
|  | Supine | HR | 72 | 4.8 | 2-6 | 66 | 4.4 | 2-5 |
|  |  | MR | 68 | 4.5 | 3-6 | 66 | 4.4 | 2-5 |
| FS | Sit | HR | 69 | 4.6 | 3-8 | 64 | 4.3 | 3-5 |
|  |  | MR | 65 | 4.3 | 3-5 | 62 | 4.3 | 2-5 |
|  | Supine | HR | 72 | 4.8 | 2-6 | 66 | 4.4 | 2-5 |
|  |  | MR | 68 | 4.5 | 3-6 | 66 | 4.4 | 2-5 |
| NO | Sit | HR | 55 | 3.7 | 1-5 | 48 | 3.2 | 1-5 |
|  |  | MR | 48 | 3.2 | 1-5 | 46 | 3.1 | 1-5 |
|  | Supine | HR | 59 | 3.9 | 2-7 | 51 | 3.4 | 1-5 |
|  |  | MR | 46 | 3.1 | 1-5 | 48 | 3.2 | 1-5 |
| RA | Sit | HR | 55 | 4.7 | 1-5 | 48 | 3.2 | 1-5 |
|  |  | MR | 48 | 3.2 | 1-5 | 46 | 3.1 | 1-5 |
|  | Supine | HR | 59 | 3.9 | 2-7 | 51 | 3.4 | 1-5 |
|  |  | MR | 46 | 3.1 | 1-5 | 48 | 3.2 | 1-5 |
| Reflex | Sit | HR | 65 | 4.3 | 2-8 | 64 | 4.3 | 3-5 |
|  |  | MR | 65 | 4.3 | 2-5 | 62 | 4.1 | 2-5 |
|  | Supine | HR | 72 | 4.8 | 2-7 | 66 | 4.4 | 2-6 |
|  |  | MR | 66 | 4.4 | 3-6 | 65 | 4.3 | 2-5 |

### Key kinematic differences between children with CP and TD children for the isometric condition

In agreement with previous studies, the first swing excursion and number of oscillations were smaller in children with CP than in TD children (figure 2a-b, table S2). However, the resting angle was only different between children with CP and TD children in the supine and not in the sitting position (figure 2c). In the sitting position (figure 2: column 1-2), the first swing excursion in children with CP was 51° smaller than in TD children (p < 0.001, CP: 62° ± 30°; TD: 113° ± 8°). Further, the number of oscillations was 2.7 smaller in children with CP compared to TD children (p < 0.001, CP: 3.7 ± 2.0; TD: 6.4 ± 1.4). Finally, the resting angle was not different between the two groups (CP: 64° ± 7°; TD: 66° ± 7°). In the supine position (figure 2: column 3-4), the first swing excursion in children with CP was 43° smaller than in TD children (p < 0.001, CP: 61° ± 29°; TD: 105° ± 8°). The number of oscillations was 1.5 smaller in children with CP compared to TD children (p < 0.05, CP: 4.2 ± 1.8; TD: 5.7 ± 1.6). In contrast to the sitting position, in the supine position the resting angle was 6° less vertical in children with CP than in TD children (p < 0.05, CP: 53° ±9°; TD: 59° ± 6°).

**Figure 2:**
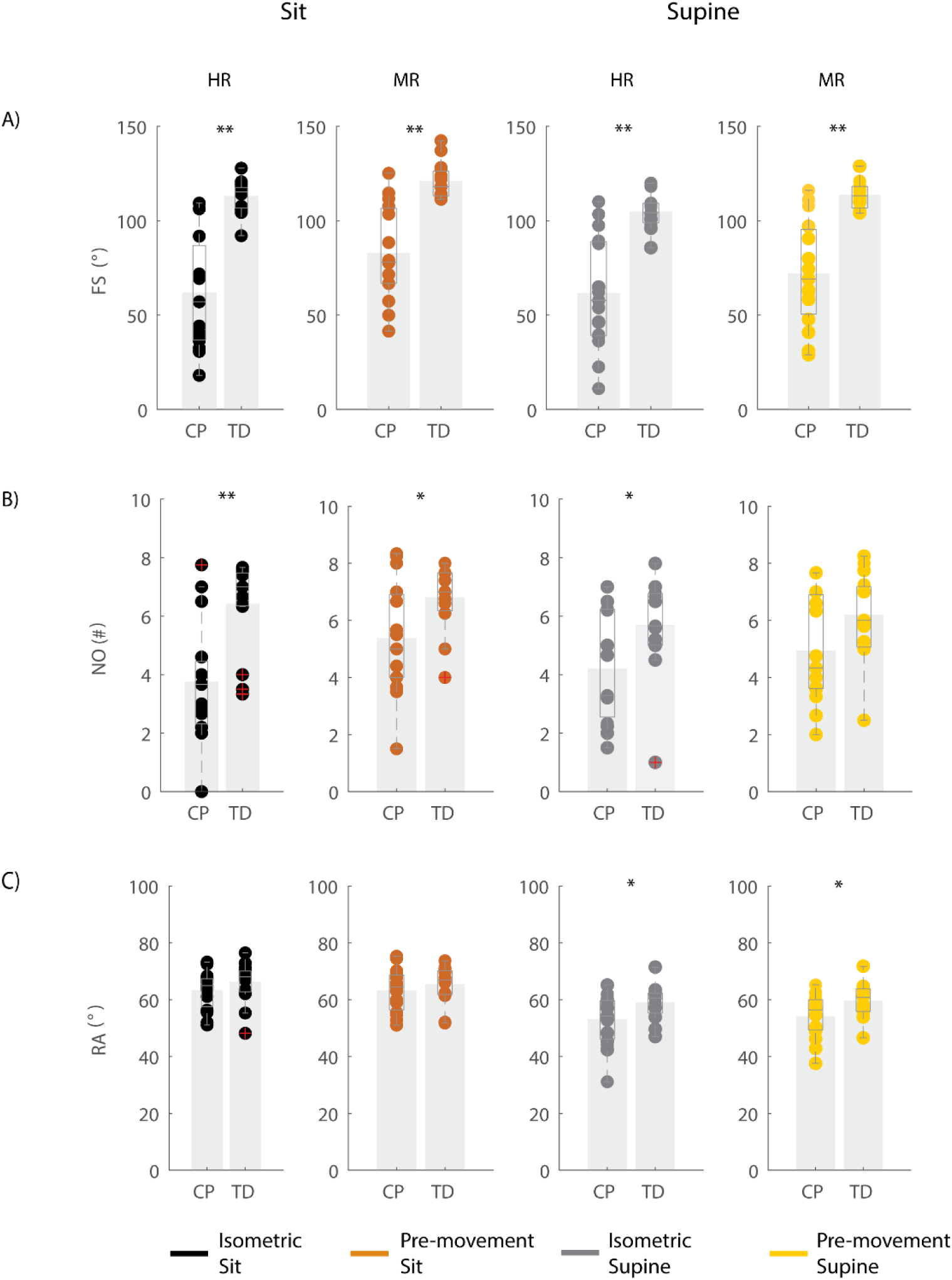
Key kinematic outcomes in children with CP and TD children. A) First swing excursion FS; B) number of oscillations NO; C) resting angle RA. Dots represent the average value over all trials for each child, light grey bars represent mean values over all children within a group. Significant differences between children with CP and TD children are indicated with * for p < 0.05 and ** for p < 0.01. Black = isometric (HR) – sitting; orange = pre-movement (MR) - sitting; grey = isometric (HR) - supine; yellow = pre-movement (MR) - supine.

### Influence of position on key kinematic parameters

The resting angle was less vertical in the supine position than in the sitting position for both groups of children (p < 0.001, CP: 64° ±7° sitting versus 53° ± 9° supine; TD: 66° ± 7° sitting versus 59° ± 6° supine), (figure 2c, table S2). Further, the first swing excursion in TD children was 8° smaller in the supine position than in the sitting position (p < 0.001, sitting: 113° ± 8°; supine: 105° ± 8°), (figure 2a). No other differences were observed between the sitting and supine position.

### Movement history alters first swing excursion and number of oscillations but not resting angle

When the leg was moved prior to release (MR), the first swing excursion and number of oscillations increased while the resting angle did not change (figure 3, 4 and 5, table S3) and the observed increases in first swing excursion and number of oscillations were larger in children with CP than in TD children. In the sitting position, the first swing excursion increased on average by 21° (p < 0.001, ± 12°) for children with CP (figure 3 and 5a) and by 8° (p < 0.001, ± 7°) for TD children (figure 4 and 5a). This increase was higher (p < 0.01) in children with CP than in TD children (figure 5a: column 1). The number of oscillations increased on average by 2 oscillations (p < 0.001, ± 1.1) for children with CP (figure 3 and 5b) and by 0.4 oscillations (p < 0.01, ± 0.4) for TD children (figure 4 and 5b). The increase in number of oscillations was higher (p < 0.001) in children with CP than in TD children (figure 5b: column 1). Movement history did not influence the resting angle (figure 5c: column 1).

**Figure 3:**
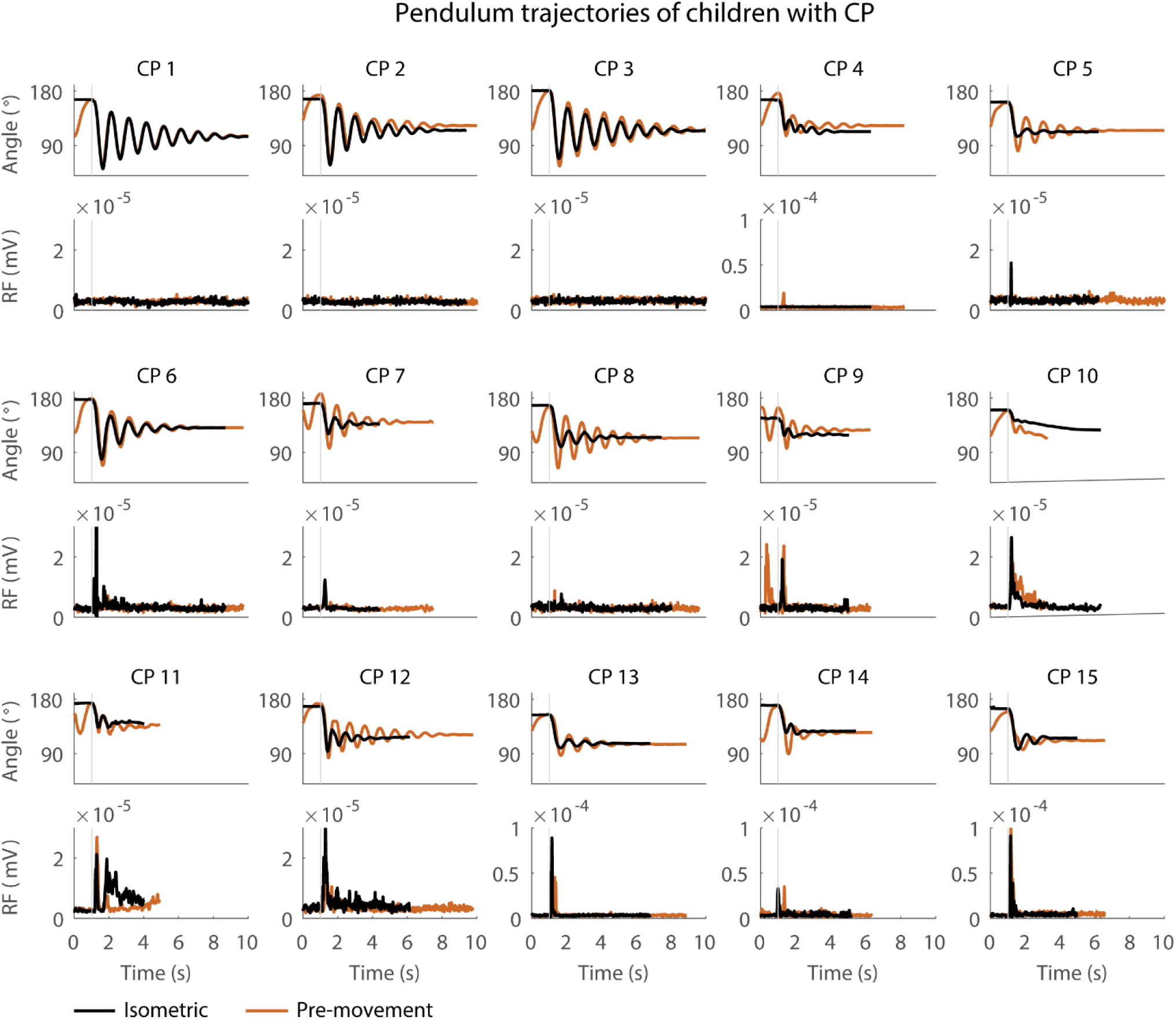
Pendulum trajectories and rectus femoris (RF) muscle activity (sEMG) of children with CP in the sitting position. A representative knee angle trajectory is shown for the isometric (black) and pre-movement (orange) conditions. Children are numbered from low reflex activity to high reflex activity based on number of trials with reflex activity (occurrence of reflexes). An increase in first swing excursion and number of oscillations is seen in the movement condition in orange as compared to the isometric condition in black.

**Figure 4:**
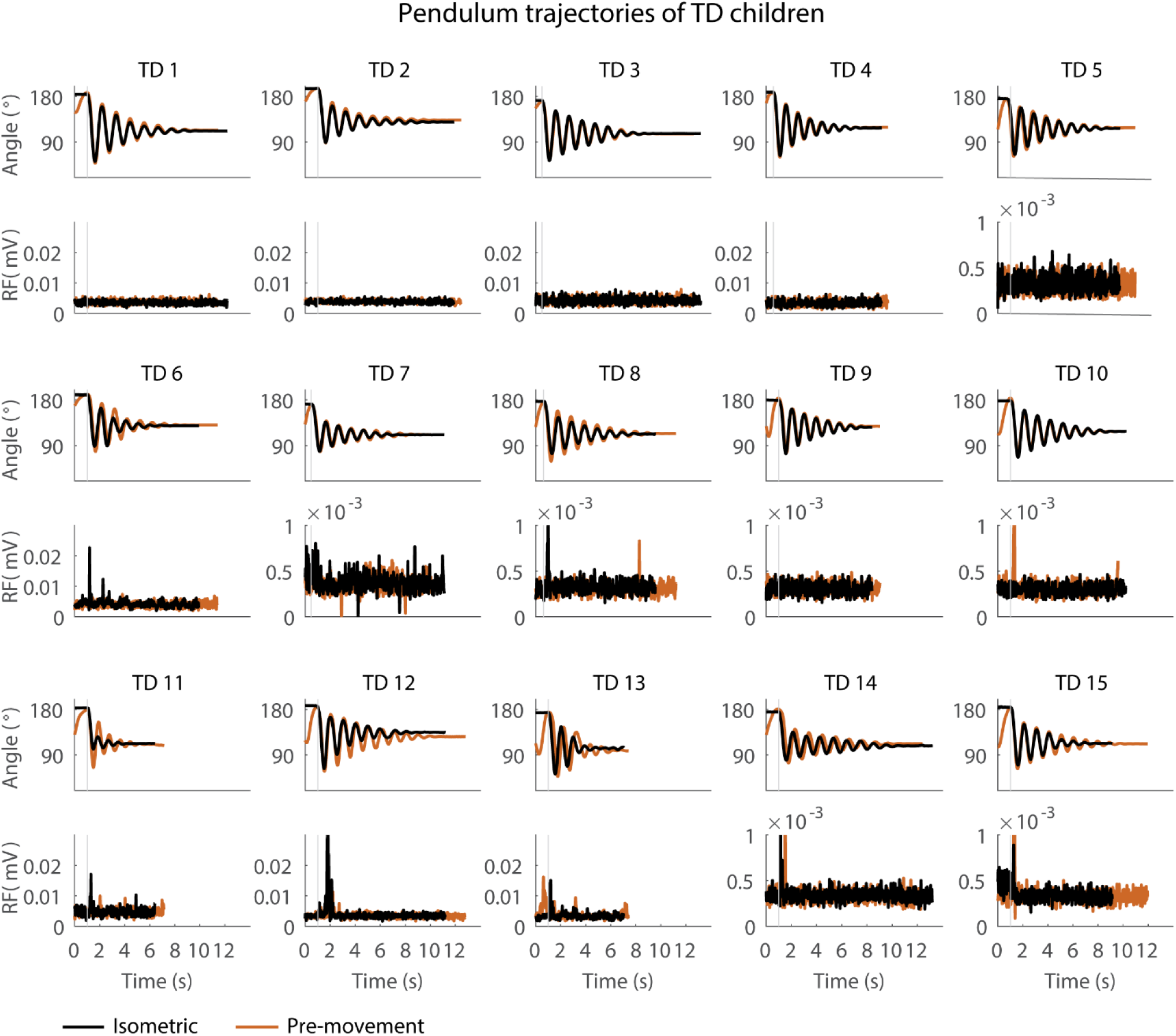
Pendulum trajectories and rectus femoris (RF) muscle activity (sEMG) of TD children in the sitting position. A representative knee angle trajectory is shown for the isometric (black) and pre-movement (orange) conditions. Children are numbered from low reflex activity to high reflex activity based on number of trials with reflex activity (occurrence of reflexes). An increase in first swing excursion and number of oscillations is seen in the movement condition in orange as compared to the isometric condition in black.

**Figure 5:**
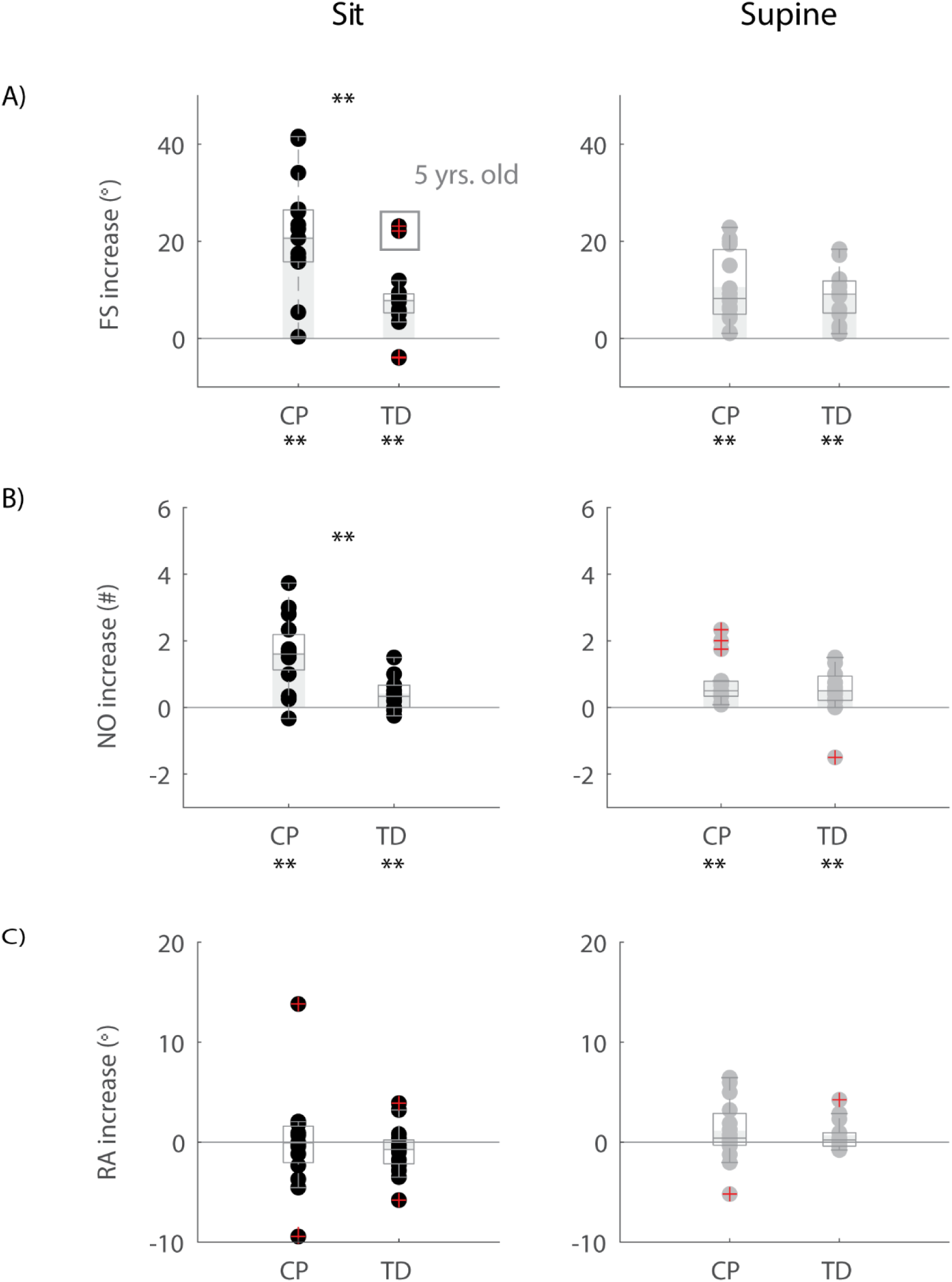
Influence of movement history on key kinematic outcomes. A) First swing excursion FS; B) number of oscillations NO; C) resting angle RA. We computed the difference in outcome parameters by subtracting outcomes in the isometric (HR) condition from outcomes in the pre-movement (MR) condition. Dots represent the average value over all trials for each child, light grey bars represent mean value over all children within a group. Significant differences between conditions are indicated with * for p < 0.05 and ** for p < 0.01 below the graph, significant differences between children with CP and TD children are indicated by * for p < 0.05 and ** for p < 0.01 above and in between graphs. Black = sitting position; grey = supine position.

Also in the supine position, the first swing excursion and number of oscillations increased with pre-movement (MR), but the increase was not different between children with CP and TD children. First swing excursion increased on average by 10° (p < 0.001, ± 7°) for children with CP and by 9° (p < 0.001, ± 5°) for TD children (figure 5a: column 2). Further, the number of oscillations increased on average by one oscillation (p < 0.01, ± 0.8) for children with CP and by 0.5 oscillations (p < 0.05, ± 0.7) for TD children (figure 5b: column 2). The increases observed in first swing excursion and number of oscillations were not different between children with CP and TD children. Also in the supine position, there was no influence of movement history on the resting angle (figure 5c: column 2).

### The effect of movement history on pendulum kinematics is more variable in children with CP than in TD children

First swing excursion was more variable in children with CP than in TD children for both the sitting and supine position (p < 0.001) (figure 2a). Also the variability in the increase in first swing excursion (p <0.01) and number of oscillations (p < 0.001) when moving the leg before releasing it was higher in children with CP than in TD children in the sitting position, but not in the supine position (figure 5a-b). Notwithstanding this high variability across children with CP, the first swing excursion and number of oscillations increased in all children with pre-movement (figure 3, figure 5a-b). Given this high variability, we explored whether a smaller first swing excursion in the isometric condition was related to a larger increase in first swing excursion in the pre-movement versus isometric condition in children with CP. However, we did not find such relation (figure S2). Children that had similar first swing excursions in the isometric condition (HR), presented different responses in the pre-movement condition (MR) (figure 3, S2).

### Higher occurrence and earlier onset of reflexes in children with CP than in TD children

We observed reflex activity more often in children with CP than in TD children during the first swing and reflexes occurred sooner after release of the leg in children with CP than in TD children (figure 6, table S4).

**Figure 6:**
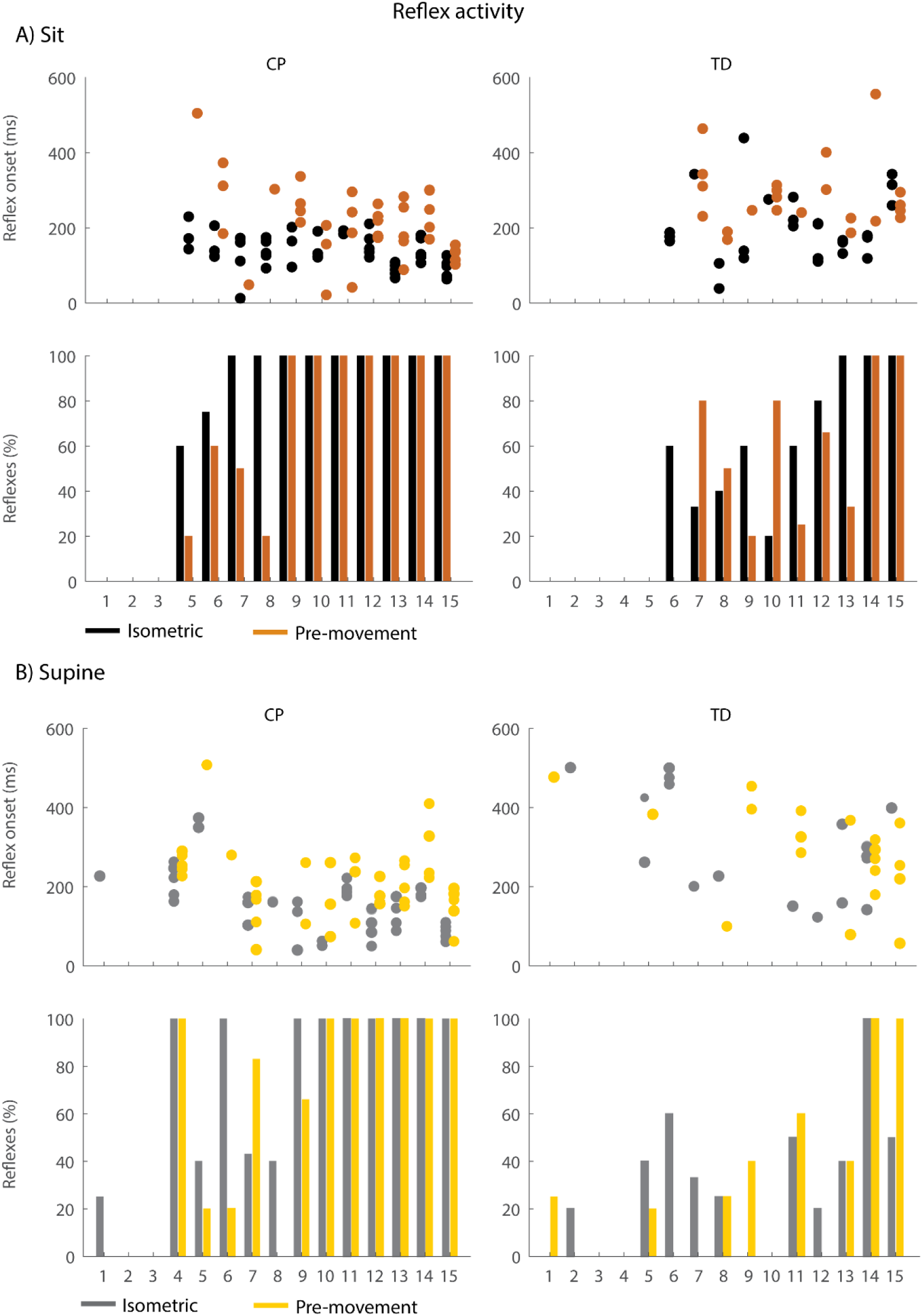
Occurrence and timing of reflex activity in the sitting (A) and supine (B) position. The dots represent reflex onset with respect to release of the leg, only for trials in which reflex activity was observed. The bars represent the percentage of trials with reflex activity. Black = isometric (HR) – sitting; orange = pre-movement (MR) – sitting; grey = isometric (HR) - supine; yellow = pre-movement (MR) – supine.

In the sitting position, the occurrence of reflex activity was higher in children with CP than in TD children (p < 0.05, CP: 73.9%; TD: 42.2%) when the leg was held isometric (HR) but not when the leg was moved prior to release (MR) (figure 6a bottom). When reflexes were observed, reflex onset was 70 ms earlier in children with CP than in TD children (p < 0.05, 142 ms ± 30 ms in CP versus 212 ms ± 78 ms in TD) in the isometric condition (HR), but reflex onset was not different between children with CP and TD children in the pre-movement condition (MR) (figure 6a top).

In the supine position, the occurrence of reflexes was higher in children with CP than in TD children in both the HR and MR conditions (p < 0.05, HR: 63.2% in CP versus 29.2% in TD; MR: 59.3% in CP versus 27.3% in TD), (figure 6b bottom). When reflexes were observed, reflex onset was 170 ms earlier in children with CP than in TD children (p < 0. 001, CP: 165 ms ± 77 ms; TD: 294 ms ± 124 ms) in the isometric condition (HR), but reflex onset was not different between children with CP and TD children in the pre-movement condition (MR) (figure 6b top).

### Movement history influences the onset of reflex activity in children with CP

Moving the leg instead of keeping it isometric prior to release did not alter the occurrence of reflex activity but delayed the onset of reflex activity by 87ms (p < 0.05) while sitting and by 78ms (p < 0.001) while supine in children with CP (figure 6: black vs. orange and grey vs. yellow, table S5). No differences in occurrence of reflex activity and in reflex onset were observed in TD children.

Movement history only altered reflex magnitude of the rectus femoris for children with CP in the sitting position. The AUC decreased when the leg was moved instead of kept isometric prior to release (p < 0.05), but we observed large variability in the responses between children with CP. The AUC decreased in the MR condition in six children, whereas it did not change in six other children and increased in two others. There was no influence of movement history on reflex magnitude for TD children (sit and supine position) and children with CP in the supine position (figure 7).

**Figure 7:**
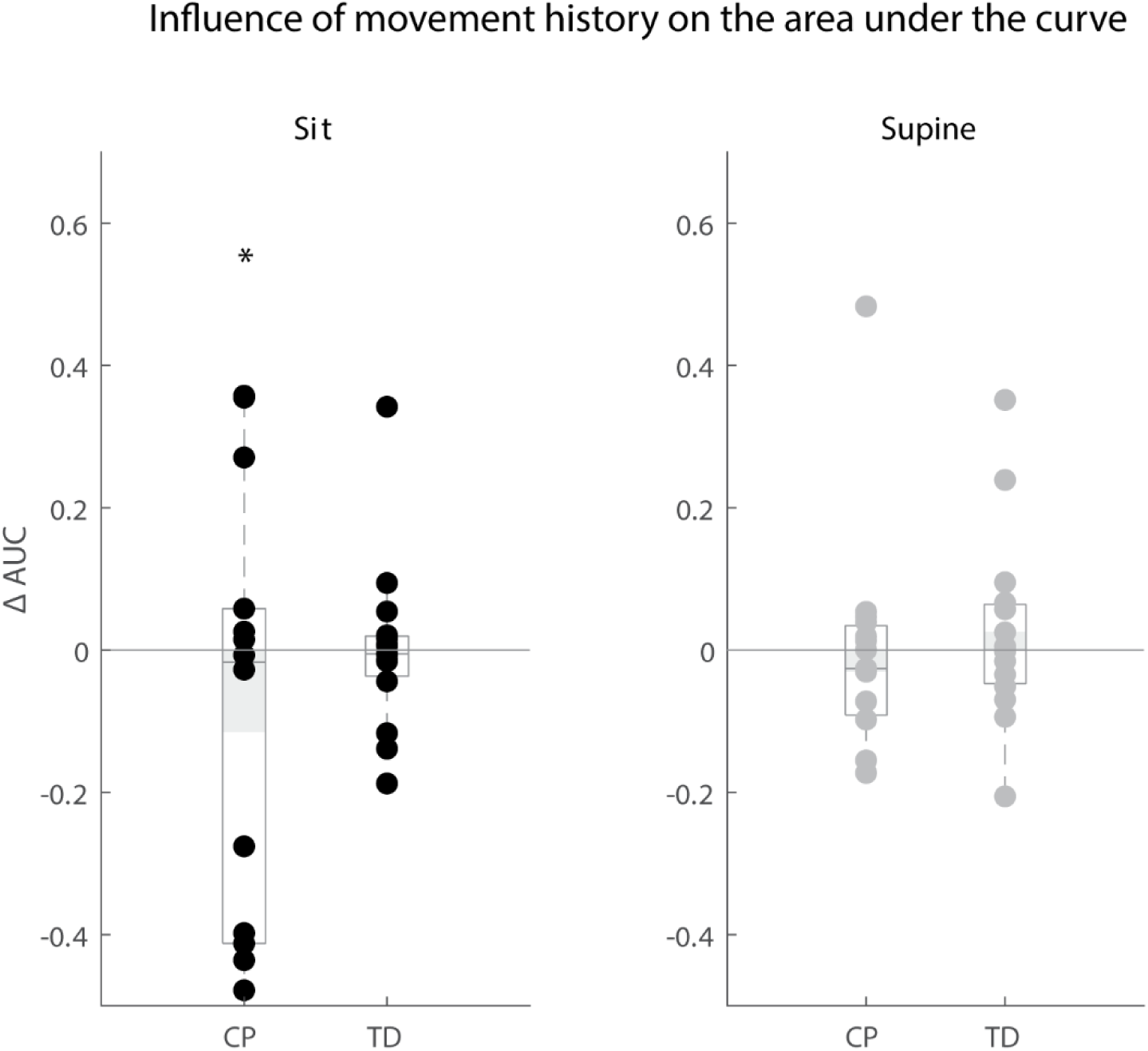
Influence of movement history on reflex magnitude. Difference in area under the curve of rectus femoris sEMG activity between the isometric and pre-movement condition. The AUC is normalized by dividing the difference between isometric and pre-movement condition by the AUC for the isometric condition. Values are presented for children with CP and TD children in black for the sitting position and in grey for the supine position. Dots represent the average value over all trials for each child, light grey bars represent the mean value over all children within a group. Significant differences are indicated with * for p < 0.05 and ** for p < 0.01.

### First swing excursion was correlated with resting angle and occurrence of reflex activity

#### Resting angle

In the supine, but not in the sitting position, children with a larger first swing excursion also had a more vertical resting angle (CP: p < 0.001, r = 0.84; TD: p < 0.01, r = 0.66), (figure 8a, table S6).

**Figure 8:**
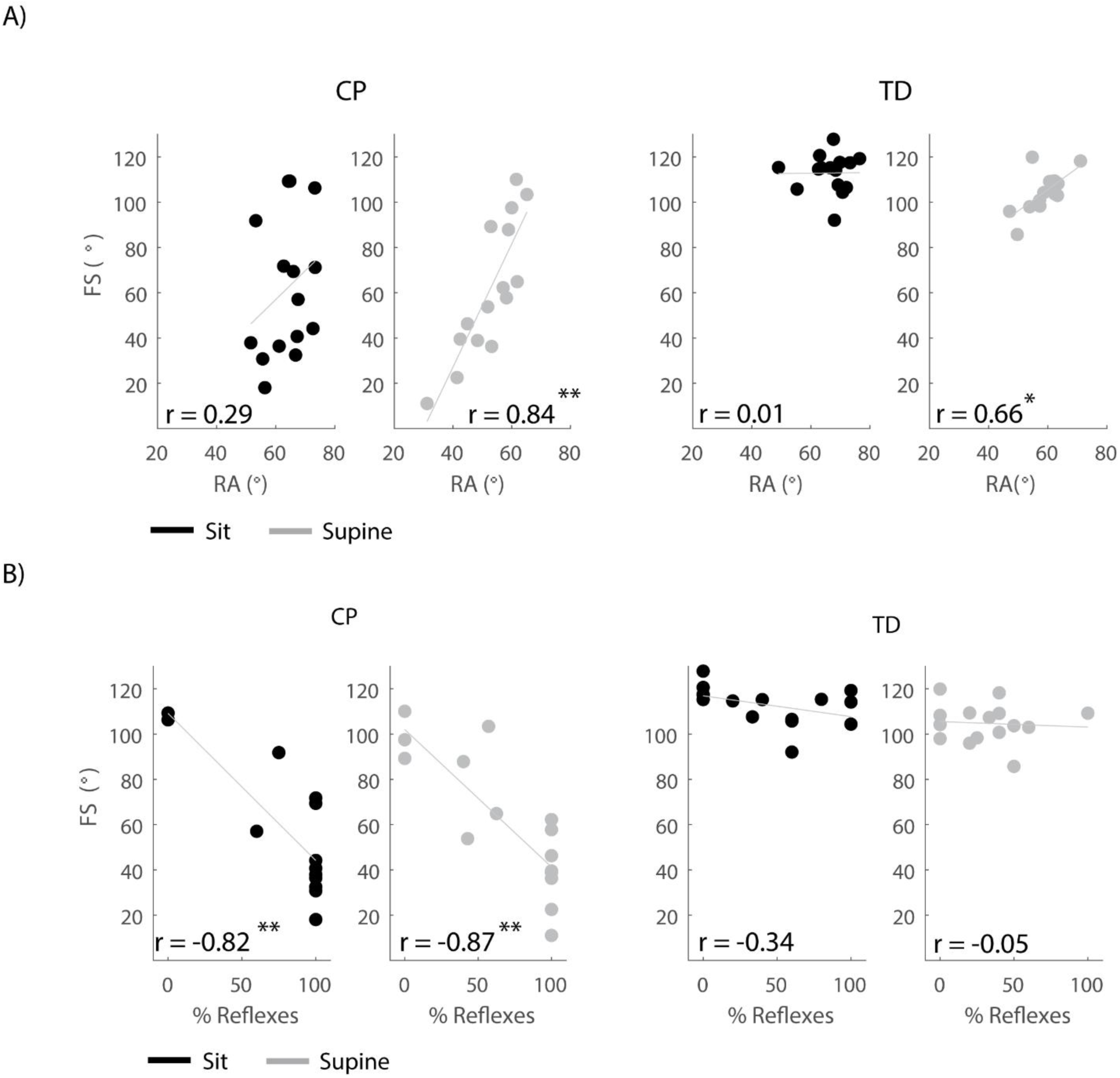
Correlations between outcome parameters. A) First swing excursion and resting angle are positively correlated in the supine position but not in the sitting position. B) First swing excursion is negatively correlated with occurrence of reflexes in children with CP but not in TD children. Significant correlations are indicated with an * for p < 0.05 and ** for p < 0.01.

#### Occurrence of reflex activity

In both the sitting and supine position, the first swing excursion was smaller in children with CP that had a higher occurrence of reflex activity (sit: p < 0.001, r = -0.82; supine: p < 0.001, r = -0.87), (figure 8b, table S6). First swing excursion and occurrence of reflex activity were not related in TD children.

## Discussion

The current study showed that movement history alters key kinematic features of the pendulum test that are related to joint hyper-resistance. When moving the leg prior to releasing it instead of keeping it still, pendulum kinematics in children with CP became more similar to pendulum kinematics in TD children. In particular, the first swing excursion and number of oscillations, two key kinematic features that are sensitive to the presence and severity of spasticity, increased (Fowler et al 2002). Further, reflex activity was delayed. As pre-movement will alter muscle short-range stiffness, our observations are consistent with the hypothesis that muscle short-range stiffness, due to its interaction with muscle tone and reflex activity, contributes to increased resistance against stretch. Our work has implications for standardizing the pendulum test as an assessment of spasticity and for understanding the role of spasticity in functional movements, where movement history differs from movement history in clinical tests.

The movement history dependence of pendulum test kinematics suggests that short-range stiffness contributes to the decreased first swing excursion and number of oscillations in individuals with spastic CP. Pre-movement decreases short-range stiffness upon stretch in isolated muscle fibers (Campbell and Moss 2002). Therefore, moving the leg prior to releasing it is expected to decrease short-range stiffness in the knee extensors, thereby reducing the resistance against movement. The increase in first swing excursion and number of oscillations when the leg was moved prior to release compared to the isometric condition in both children with CP and TD is thus in agreement with a reduced contribution from short-range stiffness.

Short-range stiffness is proportional to isometric force prior to the stretch (Campbell and Moss 2002) and therefore the high prevalence of background muscle activity in CP (Freeman Miller 2005) might explain why pre-movement had a larger effect on pendulum test kinematics in children with CP. We were not able to directly confirm the role of background muscle activity, since the magnitude of the sEMG signal also depends on the conductance of the underlying soft tissues and therefore does not provide a quantitative measure of muscle activity. Although our previous modeling study suggested that background extensor muscle activity contributes to both reduced first swing excursion and less vertical resting angle (De Groote et al. 2018), we did not find a correlation between decreased first swing excursion and less vertical resting angle in the sitting position. Possibly, this is due to other factors independently influencing first swing excursion and resting angle. For example, reflex hyper-excitability might mainly influence first swing excursion whereas background flexor activity might mainly influence resting angle, indicating that more factors should be taken into account.

We were able to test the dependency of joint hyper-resistance on active muscle force by altering the pose of the subjects. Assuming that changing the subject’s position from sitting to supine would stretch rectus femoris beyond its optimal length, we expected a decrease in active muscle force and an increase in passive force. Since short-range stiffness is proportional to active force (Blum et al. 2019; Campbell and Moss 2002), we therefore expected a smaller influence of short-range stiffness in the supine position compared to the sitting position. Indeed, we found that pre-movement had a smaller influence on first swing excursion and number of oscillations in the supine compared to the sitting position. In addition, the resting angle was less vertical in children with CP compared to TD children in the supine position but not in the sitting position, in agreement with the assumed increased contribution from passive muscle force (Gordon and Ridgway 1987; Herzog and ter Keurs 1988).

The decreased and delayed reflex activity with pre-movement suggests that the interaction between reflex hyperactivity and history-dependent short-range stiffness force contributes to joint hyper-resistance. The decrease in reflex activity with increased first swing excursion due to reduced short-range stiffness is in agreement with muscle spindles being encoders of force and yank, i.e. time derivative of force, rather than fiber length and velocity (Blum et al. 2017; Falisse et al. 2018). If muscle spindle firing would reflect changes in length and velocity, reflexes would increase and happen sooner with pre-movement due to the larger and faster muscle stretch. The interaction between short-range stiffness and reflex activity was further confirmed by the correlation between a smaller first swing excursion and a larger number of trials with reflex activity in children with CP. However, reflex activity and the influence of movement history on reflex activity varied largely across children with CP. This might be explained by differential contributions of muscle tone versus reflex activity to joint hyper-resistance in our heterogeneous group of children with CP.

Movement history dependence of joint hyper-resistance calls for standardization of movement before applying stretch in clinical tests of spasticity and suggests pre-movement as an experimental manipulation to distinguish different neural and non-neural contributions to joint hyper-resistance. Failure to standardize movement prior to muscle stretch in spasticity might introduce large variability in joint hyper-resistance, which might have contributed to the low repeatability of commonly used tests of spasticity (van den Noort et al. 2017). In agreement with literature, we found reduced first swing excursions in children with CP that have no spasticity according to MAS scores for the quadriceps muscle (table S1), confirming the higher sensitivity of the pendulum test as compared to MAS (Brown et al. 1988; Fowler et al. 2000). We believe that performing the pendulum test in both the hold and pre-movement condition might further improve assessment of spasticity. Since pre-movement affects short-range stiffness, which interacts with neural factors (e.g. background muscle activity and hyperreflexia) but not with increased resistance due to altered non-contractile tissue properties, our results suggest that the effect of pre-movement will be larger when joint hyper-resistance results from neural factors rather than from non-neural factors. Therefore, multiple test conditions might enable better discrimination between neural and non-neural contributions to joint hyper-resistance (van den Noort et al. 2017). Although the observation that first swing excursion depends on movement history was very robust across children with CP and TD children, the magnitude of the effect varied widely in our heterogeneous (based on clinical scores including MAS, GMFCS, and range of motion) group of children. A supplementary analysis showed that the variability in the effect of movement history on first swing excursion could not be explained by differences in first swing excursion in the isometric condition in children with CP (figure S3). Children with similar first swing excursions in the isometric condition responded differently to pre-movement. This suggests that differential contributions of neural and non-neural factors between subjects might contribute to the variability in the effect of movement history. Note that we also observed two outliers with higher increases in first swing excursion in the more homogenous group of TD children (Figure 5a). Remarkably, these outliers were the two youngest children, which were only 5 years old at the time of the test. Reflex excitability might be higher in TD children up to the age of six years than in older children (O’Sullivan M et al. 1991). Indeed, both children had a high occurrence of reflexes in the hold trials that decreased with pre-movement (Figure 6, TD11&13).

Assessing the effect of movement history on joint hyper-resistance might also be important to further increase our understanding of the role of spasticity in functional movements, where movement history is typically different from clinical tests. Our results suggest that joint hyper-resistance as measured during clinical tests might not be representative for the hyper-resistance during tasks requiring non-isometric muscle contractions but the discrepancy in hyper-resistance in clinical tests of spasticity and functional movements might be highly subject-specific given the high variability of the effect of movement history on first swing excursion.

Despite the fact that we tried to control the initial angle between trials and conditions, the leg was on average 1.6° (CP – max: 7.1°) and 2.1° (TD – max: 6.4°) closer to horizontal in the pre-movement than in the isometric condition. However, this difference alone cannot explain the difference in first swing excursion between conditions. We simulated lower leg kinematics based on different initial angles (De Groote et al. 2018). The simulated difference in first swing excursion due to altered initial angles was 1.5° and 4° for a difference of 2° (observed average) and 7° (observed maximum) in initial angle, respectively (figure S3). These differences in first swing excursion due to altered initial angles were both smaller than the observed differences between conditions (CP mean: 10.5°; TD mean: 8.8°). In addition, we corrected the increase in first swing excursion with the simulated difference in first swing excursion due to the difference in initial angle and repeated the statistical analysis based on the corrected values. Our results and conclusions did not change.

## Conclusion

Our study supports a novel hypothesis about the mechanism of joint hyper-resistance derived from a previously published computational model of the pendulum test of spasticity. The movement history dependence of pendulum kinematics and reflex activity supports our hypothesis that muscle short-range stiffness and its interaction with increased background muscle activity and reflex hyper-excitability contribute to joint hyper-resistance in spastic CP. Our results suggest that movement history should be taken into account when assessing joint hyper-resistance and when studying its contribution to movement impairments. We demonstrate that the period of rest before stretching the muscle in clinical tests of spasticity is critical for the following response. Therefore, the period of rest before imposing the stretch should be standardized. In addition, the response to muscle stretch in a relaxed patient might be very different from the response to muscle stretch in the same patient performing functional movements as both movement history and background muscle activity will be different in both conditions. Assessing the effect of movement history in clinical tests of spasticity might be a first step towards a better understanding of the role of joint hyper-resistance in movement impairments. Our results suggest that there might be large variability in the effect of pre-movement between patients that present with the same clinical score, which might necessitate a more personalized treatment of hyper-resistance problems.

## Data Availability

Data will be available upon request to the authors and upon publication

## List of abbreviations

AUC: Area under the curve
CP: Cerebral palsy
FS: First swing excursion
GMFCS: Gross motor function classification scale
HR: Hold and release
IA: Initial angle
MAS: Modified Ashworth scale
MR: Movement and release
NO: Number of oscillations
RA: Resting angle
RF: Rectus femoris
sEMG: Surface electromyography
TD: Typically developing

## Author Contributions

Conceptualization: LT and FD. Recruitment: JW and AV. Data collection and curation: JW. Methodology: JW, LT, FD. Formal analysis and visualization: JW and FD. Funding acquisition: JW, LT, FD. All authors contributed to interpretation of the results and in writing the manuscript.

## Acknowledgments

We would like to thank Sistania Bong for designing the back rest and all our participants for participating in this study.

## Supplementary material

**Figure S1:**
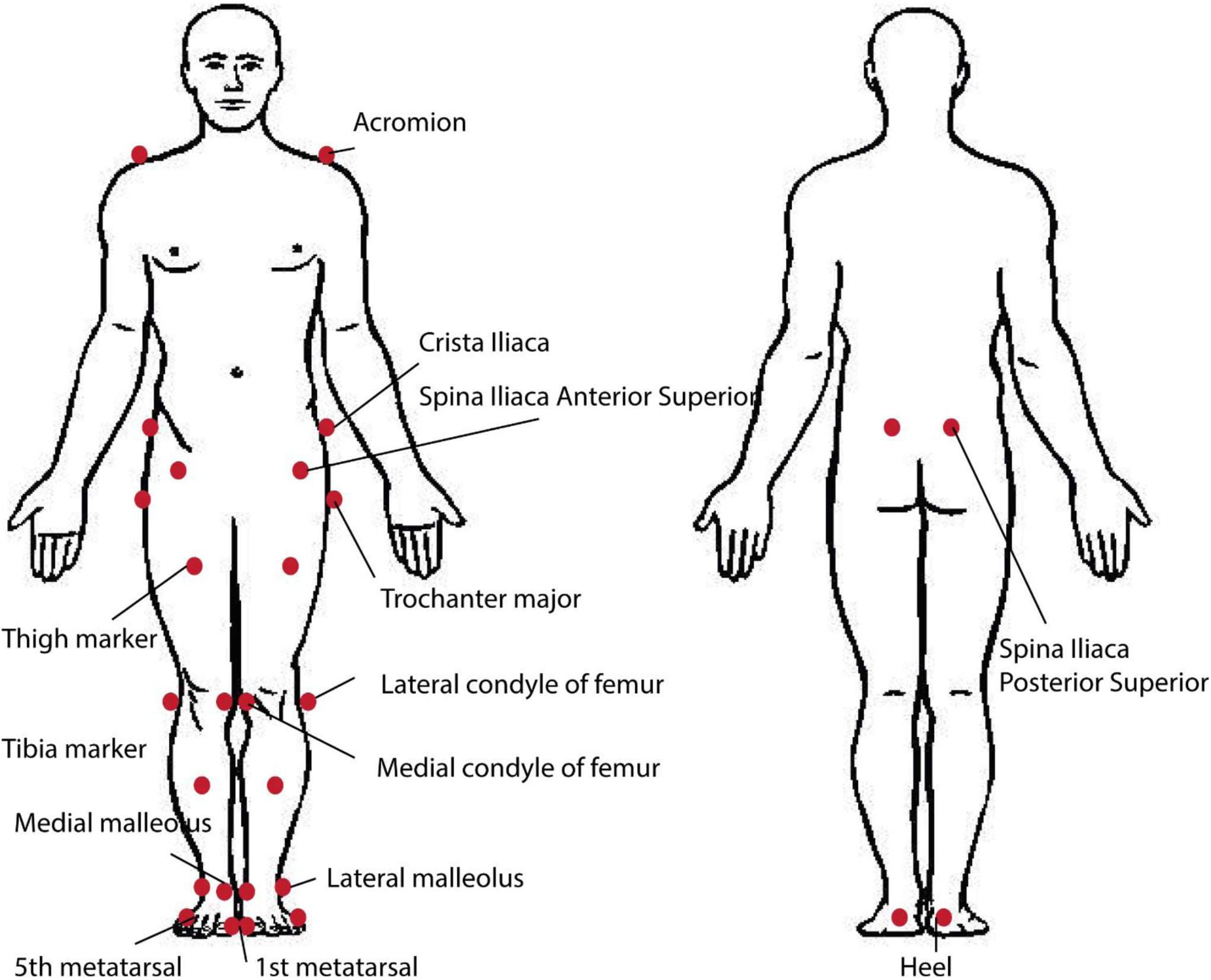
Marker set-up used to measure kinematics during the pendulum test in children with CP and TD children.

**Figure S2:**
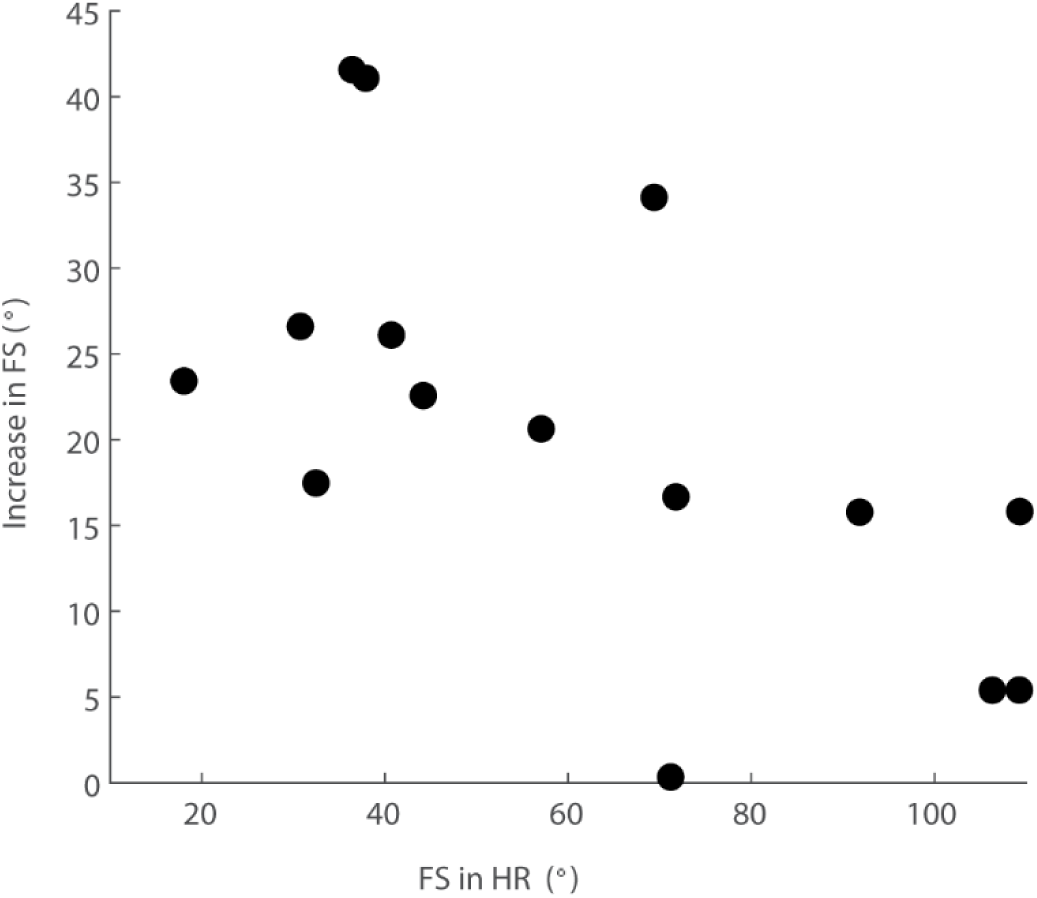
Relation between first swing excursion in the isometric condition and the increase in first swing excursion in the pre-movement with respect to the isometric condition for children with CP in the sitting position.

**Figure S3:**
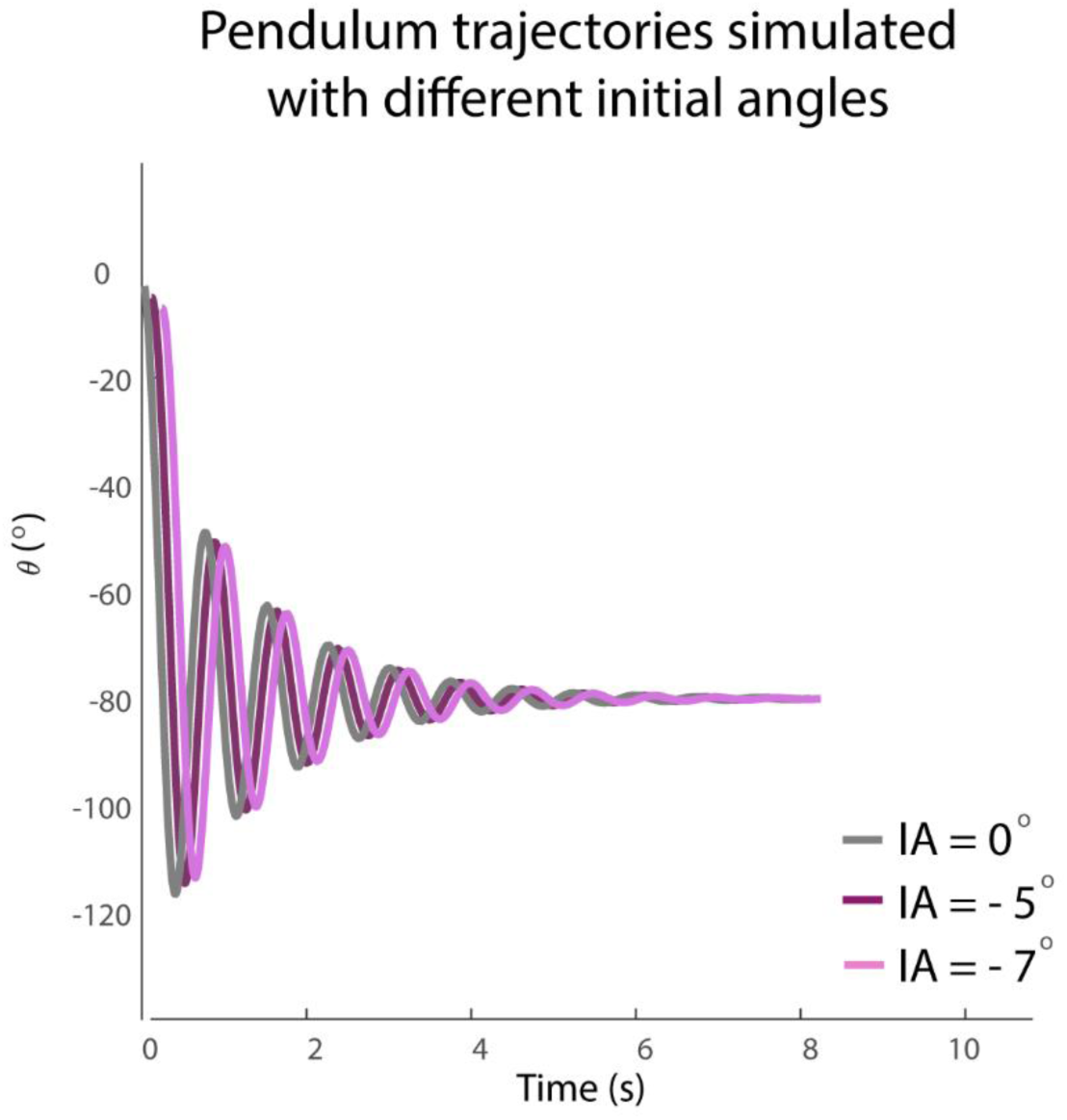
Simulated pendulum movement for three different initial angles: 0° (grey), -5° (purple), -7° (pink). Only minor differences are observed in the first swing excursion between trials. Curves are shifted horizontally for clarity. Kinematic trajectories were simulated based on the model described by De Groote et al (2018). The model with short-range stiffness, but without reflex activity was used, since the largest differences based on initial angle were expected in this configuration. Muscle tone was set on 0.4 Nm. We used values for mass (2.4kg), length (0.26m) and inertia (0.21kg/m^2^) that were representative for the subjects in our database.

**Table S1:**
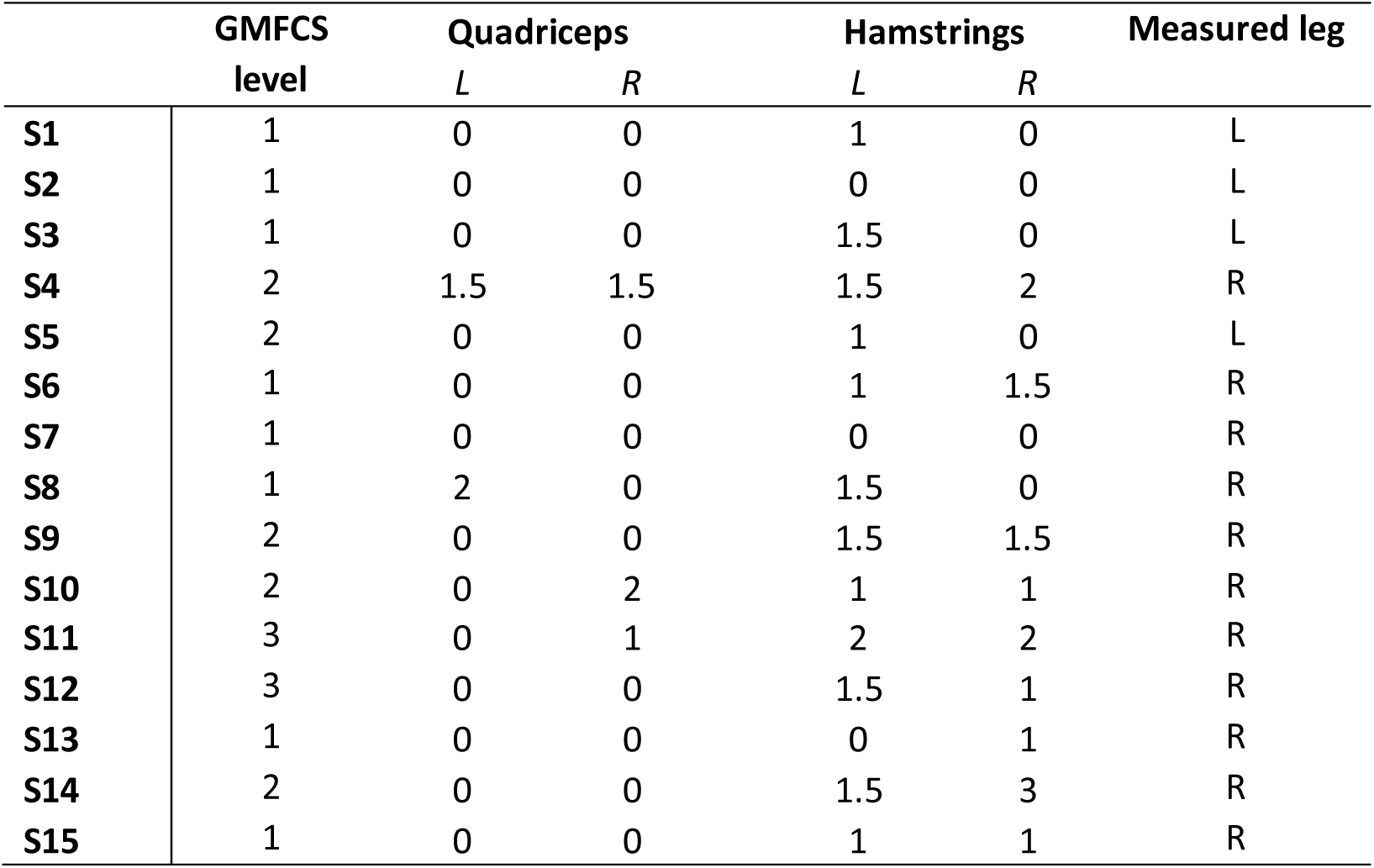
Spasticity scores as measured by the Modified Ashworth Scale for children with CP

**Table S2:** a) Key kinematic outcomes (mean and standard deviation). b) p-values for the comparisons between subject groups (CP and TD) and positions (sitting and supine).

| a) | Sit (HR) |  |  |  | Supine (HR) |  |  |  |
| --- | --- | --- | --- | --- | --- | --- | --- | --- |
|  | CP |  | TD |  | CP |  | TD |  |
|  | *Mean* | *SD* | *Mean* | *SD* | *Mean* | *SD* | *Mean* | *SD* |
| **FS (°)** | 62 | 30 | 113 | 8 | 61 | 29 | 105 | 8 |
| **NO (#)** | 3.7 | 2.0 | 6.4 | 1.4 | 4.2 | 1.8 | 5.7 | 1.6 |
| **RA (°)** | 64 | 7 | 66 | 7 | 53 | 9 | 59 | 6 |

| b) | CP vs. TD |  | Sit vs. Supine |  |
| --- | --- | --- | --- | --- |
|  | Sit | Supine | CP | TD |
| **FS** | < 0.001 | < 0.001 | 0.90 | <0.001 |
| **NO** | < 0.001 | < 0.05 | 0.06 | 0.07 |
| **RA** | 0.32 | < 0.05 | <0.001 | <0.001 |

**Table S3:** a) Influence of movement history on key kinematic outcomes (mean and standard deviation). b) p-values for the comparison between conditions (isometric vs. pre-movement). c) p-values for the comparison of the influence of pre-movement on the key kinematic outcomes between groups (CP vs TD) for the outcomes that were influenced by pre-movement.

| a) | Sit |  |  |  | Supine |  |  |  |
| --- | --- | --- | --- | --- | --- | --- | --- | --- |
|  | CP |  | TD |  | CP |  | TD |  |
|  | *Mean* | *SD* | *Mean* | *SD* | *Mean* | *SD* | *Mean* | *SD* |
| **Fs (°)** | 21 | 12 | 8 | 7 | 10 | 7 | 9 | 5 |
| **No (#)** | 1.7 | 1.1 | 0.4 | 0.4 | 0.7 | 0.8 | 0.5 | 0.7 |
| **Ra (°)** | 64 | 6 | 66 | 7 | 3 | 8 | 59 | 6 |

| b) | Sit |  | Supine |  |
| --- | --- | --- | --- | --- |
|  | CP | TD | CP | TD |
| **Fs** | < 0.001 | < 0.001 | < 0.001 | < 0.001 |
| **No** | <0.001 | < 0.01 | < 0.01 | < 0.05 |
| **Ra** | 0.98 | 0.26 | 0.11 | 0.27 |

| c) | CP vs TD |  | Sit vs. Supine |  |
| --- | --- | --- | --- | --- |
|  | Sit | Supine | CP | TD |
| **Fs** | < 0.005 | 0.47 | < 0.01 | 0.66 |
| **No** | < 0.001 | 0.45 | < 0.01 | 0.68 |

**Table S4:** a) EMG-based outcomes describing rectus femoris reflex activity (occurrence of reflex activity (%), onset of reflexes (ms) AUC, mean and standard deviation). b) p-values for the comparisons between subject groups (CP and TD) and positions (sitting and supine).

| a) | Sit (HR) |  |  |  | Supine (HR) |  |  |  |
| --- | --- | --- | --- | --- | --- | --- | --- | --- |
|  | CP |  | TD |  | CP |  | TD |  |
|  | *Mean* | *SD* | *Mean* | *SD* | *Mean* | *SD* | *Mean* | *SD* |
| **Occurrence (%)** | 74 | 40 | 42 | 38 | 63 | 41 | 29 | 27 |
| **Reflex onset (ms)** | 142 | 30 | 212 | 78 | 165 | 77 | 294 | 124 |
| **AUC** | 0.0044 | 0.0029 | 0.0013 | 0.0010 | 0.0035 | 0.0021 | 0.0012 | 0.0009 |

Movement history influences pendulum kinematics
| b) | CP vs. TD |  | Sit vs. Supine |  |
| --- | --- | --- | --- | --- |
|  | Sit | Supine | CP | TD |
| **Occurrence (%)** | < 0.05 | < 0.05 | 0.11 | 0.16 |
| **Reflex onset (ms)** | < 0.05 | < 0.001 | 0.56 | 0.05 |
| **AUC** | < 0.001 | < 0.001 | 0.17 | 0.17 |

**Table S5:**
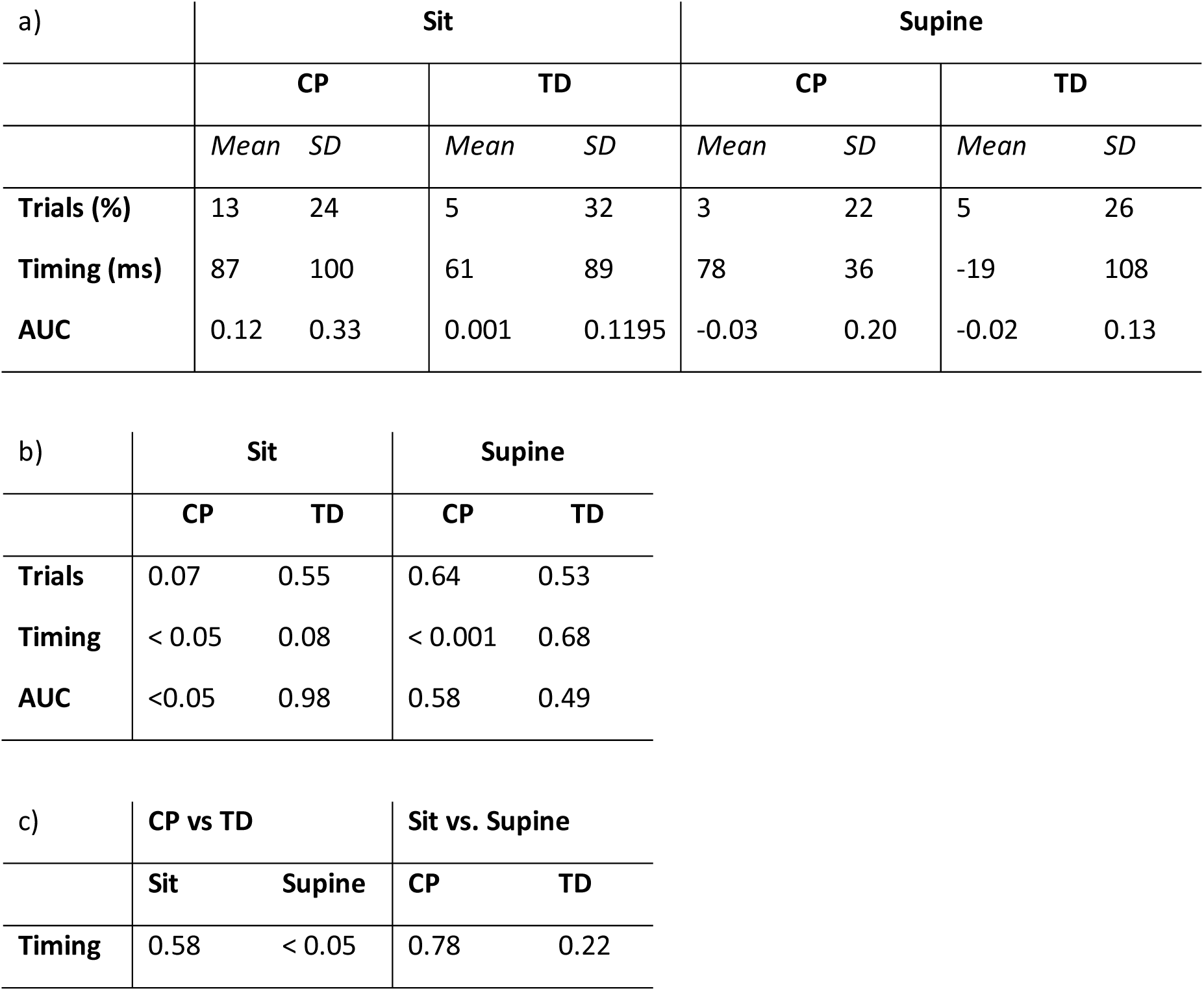
a) Influence of movement history on EMG-based outcomes describing rectus femoris reflex activity (mean and standard deviations). b) p-values for the comparison between conditions (isometric and pre-movement). c) p-values for the comparison between groups for parameters that were influenced by pre-movement.

**Table S6:** a) Correlations between first swing excursion and resting angle in the isometric (HR) condition. b) Correlations between first swing excursion and occurrence of reflexes in the isometric (HR) condition.

| a) | CP |  | TD |  |
| --- | --- | --- | --- | --- |
|  | *r* | *p* | *r* | *p* |
| Sit | 0.29 | 0.30 | 0.01 | 0.97 |
| Supine | 0.84 | <0.001 | 0.66 | < 0.01 |

| b) | CP |  | TD |  |
| --- | --- | --- | --- | --- |
|  | *r* | *p* | *r* | *p* |
| Sit | - 0.82 | < 0.01 | - 0.34 | 0.21 |
| Supine | - 0.87 | < 0.01 | - 0.05 | 0.86 |

